# The prevalence of chronic kidney disease in people with severe mental illness: A systematic review protocol

**DOI:** 10.1101/2024.09.04.24313090

**Authors:** C. Carswell, K. Bramham, J. Chilcot, R. Jacobs, D. Osborn, N. Siddiqi

**Affiliations:** Department of Health Sciences, University of York, York, UK; King’s College Hospital NHS Trust, London, UK; Institute of Psychiatry, Psychology and Neuroscience, King’s College London, London, UK; Centre for Health Economics, University of York, York, UK; Division of Psychiatry, University College London, London, United Kingdom; Hull York Medical School, York, UK; Bradford District Care NHS Foundation Trust, Bradford, UK

## Abstract

**Background:** People with severe mental illness (SMI) are more likely to develop long-term physical health conditions, including type 2 diabetes and cardiovascular disease, compared to people without SMI. This contributes to an inequality in life expectancy known as the ‘mortality gap’. Chronic kidney disease (CKD) is a growing global health concern set to be the 5th leading cause of life-years lost by 2040. However, there is limited research exploring the relationship between CKD and SMI. This systematic review will aim to examine the prevalence and incidence of CKD among people with SMI.

**Methods:** We will search Medline, Embase, PsycINFO, CINAHL, Scopus and Web of Science for primary epidemiological research reporting the prevalence or incidence of CKD among people with SMI in any setting. Retrieved records will be managed in Covidence and screened by two independent reviewers. Data will be extracted from included studies using a piloted data extraction form, and the quality of studies will be evaluated using the appropriate JBI Critical Appraisal Checklist. The certainty of evidence will be assessed using the Grading of Recommendations, Assessment, Development, and Evaluations (GRADE) approach. Data from the included studies will be narratively synthesised. Meta-analyses will be conducted using random effects models to examine the prevalence and incidence of CKD among people with SMI.

**Discussion:** There is limited research exploring the relationship between CKD and SMI, and this proposed systematic review will be the first to examine the prevalence of CKD among people with SMI. This review will highlight the extent of the problem and provide a foundation for future research to improve health outcomes for people with SMI.

## Introduction

People with severe mental illness (SMI; enduring conditions that can present with psychosis, including schizophrenia and bipolar disorder) die, on average, 15-20 years earlier than people who do not have SMI [1, 2]. This inequality, known as the mortality gap, is widening over time and is largely driven by higher rates of long-term physical health conditions and associated poor outcomes [1, 3]. The leading cause of death among people with SMI is cardiovascular disease, accounting for 70% of all deaths in people with either bipolar disorder or schizophrenia [4]. The risk of sudden cardiac death or cardiovascular mortality is five times higher in people with SMI, compared to people without SMI [4].

While research has explored the relationship between SMI and long-term conditions such as cardiovascular disease [5] and type 2 diabetes [6], chronic kidney disease (CKD) has not received the same attention [7]. CKD is a condition characterised by progressive loss of kidney function that can eventually lead to kidney failure (CKD stage 5), requiring kidney replacement therapies such as a transplant or dialysis [8]. The current global estimate for the prevalence of CKD is 843.6 million [9]. However, prevalence is increasing, and CKD is set to become the 5th leading cause of life-years lost globally by 2040 [10].

There is evidence to suggest that people living with SMI could be at higher risk of developing CKD [7]. Antipsychotic medication and mood stabilisers, commonly used in the treatment of SMI, can induce metabolic syndrome, and increase the risk of diabetes mellitus and hypertension which are the leading causes of CKD worldwide [11]. Lithium, a common mood stabiliser used in the treatment of bipolar disorder, is nephrotoxic [12], can induce acute kidney injury at high doses, and increases the risk of CKD with long-term use [13]. Antipsychotic medications may also increase the risk of developing CKD [14]. Although not known to be directly nephrotoxic, this risk likely results from the significant cardiometabolic disturbances that can result as a side-effect of antipsychotic medication [15], and the well-established relationship between CKD and poor cardiometabolic health [16]. In addition, lifestyle and behavioural factors such as smoking [17], diets high in fat, salt and sugar [18], and high levels of sedentary behaviour are known risk factors for CKD, while also being more common among people with SMI [19-22].

Identifying people at risk of CKD is crucial to facilitate early identification and intervention [23]. Early intervention can prevent or slow progression to the later stages of CKD, and reduce the risk of cardiovascular mortality [24]. This is pertinent as cardiovascular complications are the leading cause of death related to both CKD and SMI [25, 26], yet research suggests early intervention is not occurring in this population. For example, people with SMI have lower rates of accessing nephrology care [27], are less likely to receive dialysis [28, 29], and are less likely to be assessed for transplantation when compared to people without SMI [30].

To advocate for improved identification and appropriate care for people with co-existing SMI and CKD, there is a need to understand the epidemiology of this co-morbidity, including prevalence and incidence. Therefore, a systematic review is needed to understand the extent of the problem and provide a foundation for future research.

### Objectives

This review has two overarching objectives:

- Examine the prevalence and incidence of CKD among people with SMI.
- Compare the prevalence and incidence of CKD among people with SMI, to those who do not have SMI.

## Material and Methods

This protocol has been prospectively registered on the PROSPERO database (ID: CRD42024527215) [31] and is reported in line with the Preferred Reporting Items for Systematic Reviews and Meta-Analysis Protocols (PRISMA-P) statement [32].

### Search strategy and information sources

We will search electronic databases, including Medline, Embase, PsycInfo, CINAHL, Scopus and Web of Science from conception to June 2024. No restrictions on year of publication or publication status will be applied during the initial searches, to enable exploration of prevalence and incidence across time. All searches will be re-run before the final analysis to ensure all relevant publications are included. Search strings for electronic databases were developed according to the exposure (severe mental illness), outcome (chronic kidney disease) and study design (epidemiological designs). Search terms were identified from relevant systematic reviews capturing similar concepts and were refined through initial piloting and consultation with subject librarians. The full MEDLINE strategy can be found in Appendix 1.

We will review reference lists of included articles, and seminal publications and search key journals in the subject area to ensure all relevant publications have been identified. This will include the Clinical Journal of the American Society of Nephrology, the International Journal of Nephrology, JAMA Psychiatry, The Lancet Psychiatry, BMC Nephrology and BMC Psychiatry. We will also search grey literature repositories including ProQuest Dissertations and Open Science Framework (OSF). Experts in the field of severe mental illness and kidney disease, and authors of relevant studies, will be consulted to identify any potential key publications that could have been missed in the initial searches.

### Eligibility criteria

### Study design

#### Inclusion criteria

- Epidemiological observational studies, including cohort, case-control and cross-sectional studies will be included.

#### Exclusion criteria

- Qualitative studies, randomised controlled trials, and quasi-experimental studies will be excluded. While experimental studies may report the proportions of participants with certain conditions, they are not designed to determine the prevalence or incidence of a condition and often include super-selected samples which may not be representative of the population. Therefore, they will be excluded.
- Case reports, editorials, commentaries, and protocols will be excluded.

Population/ Exposure

#### Inclusion criteria

- Adults aged 18 years and over
- The population includes participants who have a diagnosis of SMI. SMI will be defined in this review as psychiatric conditions that can present with psychosis (not induced by substances or caused by an organic condition) [33-36]. These conditions include schizophrenia, schizoaffective disorder, bipolar disorder, severe depression with psychosis, other specified psychosis, and persistent delusional disorders. Any report of participants having a diagnosis of these conditions, using any standardised diagnostic tool (including the International Classification of Diseases (ICD) and the Diagnostic and Statistical Manual (DSM)) will be included.

#### Exclusion criteria

- Participants under the age of 18. If the study has also collected data from adults (over and those under 18, it will be included if the data is presented separately and can be analysed separately.
- Studies which focus exclusively on conditions which do not meet the classification for SMI, for example, anxiety disorders, depressive disorders which do not present with psychosis, eating disorders, and personality disorders, will be excluded. Studies focused on depressive disorders, in general, will only be included if they report results for severe depression with psychosis separately. Studies which focus on a variety of mental health conditions will be included if they report findings in a way that allows identification and separate analysis of participants with SMI.
- Studies, where different mental health diagnoses or categories are not reported separately (for example, where results for participants with SMI are aggregated with participants who have common mental disorders, such as depression or anxiety), will only be included if more than 50% of the sample are known to have SMI.
- Studies where the proportion of participants with SMI cannot be determined will be excluded.

### Outcome

### Inclusion criteria

- Studies which report the prevalence or incidence of CKD, at any stage, among people with SMI.
- Studies which compare the prevalence or incidence of CKD, at any stage, among the general population (or those without SMI) to people with SMI.

### Exclusion criteria

- Studies which focus on the prevalence or incidence of acute kidney injury (AKI), without reporting the prevalence or incidence of CKD, among people with SMI.
- Studies which report the prevalence of SMI among people with CKD (where a population with CKD is used as the denominator population or exposure, and SMI is reported as the outcome).

### Study selection

Records will be imported into Covidence for screening [37]. Two independent reviewers will complete the title and abstract screening, and disagreements will be resolved through discussion. If an agreement cannot be reached, the decision will be made by a third reviewer. Following title and abstract screening the full texts of eligible studies will be imported into Covidence to undergo full-text screening, which again will be completed by two independent reviewers, with disagreements resolved through discussion or a decision from a third reviewer.

### Data extraction

Two independent reviewers will perform the data extraction. A data extraction form will be developed in Excel and initially piloted by the two reviewers on a subset of included studies to ensure the form is fit for purpose. Disagreements and inconsistencies in data extraction will be reviewed and discussed, and the initial data extraction form will be refined as needed. Following the piloting, data extraction will be performed again by two reviewers, with any disagreements being resolved by a third reviewer who has not previously been involved with the data extraction process. Articles reporting data from the same study in different publications will be grouped at this stage and presented within a single study. The data extraction form will capture the author’s names, date of publication(s), publication type(s), country, study design, total sample size, number of participants with SMI, diagnoses, mean age, percentage of females, ethnicity, medication use, data collection procedures, statistical approach, summary results (prevalence, incidence and comparisons), and covariates considered.

### Risk of bias assessment

Two independent reviewers will use the appropriate JBI (formerly Joanna Briggs Institute) Critical Appraisal Checklist to evaluate the risk of bias in each of the included studies. The JBI Critical Appraisal Checklists assess the trustworthiness and quality of published research and include checklists for a range of different methodologies including cohort studies, case-control studies and cross-sectional studies [38]. Any disagreement in the assessment of the two independent reviewers will be settled through discussion, or if an agreement cannot be reached, through a decision made by a third reviewer who has not previously been involved with the critical appraisal process.

### Certainty of evidence

We will use the Grading of Recommendations, Assessment, Development, and Evaluations (GRADE) approach to determine the certainty of the evidence [39]. GRADE outlines five domains that are assessed to determine certainty: risk of bias, inconsistency, indirectness, imprecision and publication bias. GRADE is predominantly used to assess certainty in intervention research and has not been formally adapted for the use in systematic reviews of prevalence or incidence. However, previous systematic reviews have used GRADE to evaluate the certainty of prevalence studies with adapted components [40]. We will use the GRADE guidance for baseline risk or overall prognosis, as recommended by Borges Migliavaca et al. (2020) for systematic reviews of prevalence [41].

### Data synthesis

The data from the included studies will be presented in tables and narratively synthesised to provide a summary of the study characteristics and findings. Meta-analyses will also be conducted using random effects models assuming an adequate number of studies (at least two) and clinical similarity to justify the pooling of prevalence or incidence rate. The heterogeneity across studies will be assessed through visual inspection of forest plots, a chi-squared test for heterogeneity, and the I^2^ statistic. However, as meta-analyses of prevalence tend to have high I^2^ statistics [42], we will not apply statistical thresholds to determine whether it is appropriate to pool the data. We will transform the raw proportions using the Freeman-Tukey variant of the arcsine square root transformation [43]. For objective 1, the pooled prevalence or incidence rate of CKD among people with SMI will be presented with 95% CIs. For objective 2, we will undertake a meta-analysis of prevalence or incidence rate comparisons if the data is available. These will be presented as pooled risk ratios or odds ratios, with 95% CIs. If there is sufficient data, we will conduct subgroup analyses to explore the differences in CKD across different types of SMI diagnosis, CKD stage, medication, gender, age, geography (Low- and middle-income countries or high-income countries), setting (communtiy or inpatient), and year of data collection.

## Discussion

This proposed review aims to address a crucial gap in the evidence base by synthesising the literature on the prevalence of CKD among people with SMI. Previous systematic reviews have established that people with SMI are at significantly higher risk of developing long-term physical health conditions, including type 2 diabetes [5, 6]. However, the available literature on the prevalence or incidence of CKD has not yet been synthesised in a systematic review.

### Implications for research, practice or policy

By examining the prevalence of CKD in people with SMI, this review will highlight the need for future research and policy change. Most research to date in this population has focused on the role of lithium as a risk factor for CKD [44], and there is limited research exploring other, potentially modifiable, risk factors [7]. Establishing the prevalence of CKD among people with SMI (including those not on lithium treatment) is an important step towards identifying key modifiable risk factors that contribute to the development and progression of CKD in this population. This will enable not just more targeted approaches to screening and monitoring to facilitate early intervention but could also inform the development of future interventions to improve CKD outcomes for people with SMI.

## Data Availability

No datasets were generated or analysed during the current study. All relevant data from this study will be made available upon study completion.

## Appendix 1

1. Exp Renal Dialysis/
2. h?emodialysis.tw,kf.
3. h?emofiltration.tw,kf.
4. h?emodiafiltration.tw,kf.
5. dialysis.tw,kf.
6. (Peritoneal adj1 (dialysis)).tw,kf.
7. (PD or CAPD or CCPD or APD).tw,kf.
8. Exp Renal Insufficiency/
9. Exp Kidney Failure/
10. Exp Renal Insufficiency, Chronic/
11. Exp Kidney Diseases/
12. Uremia/
13. (endstage adj1 (renal or kidney)).tw,kf
14. (ESRF or ESKF or ESRD or ESKD).tw,kf
15. (chronic adj1 (kidney or renal)).tw,kf
16. (CKF or CKD or CRF or CRD).tw,kf.
17. (predialysis).tw,kf.
18. (renal adj1 (transplant* or graft)).tw,kf.
19. (kidney adj1 (transplant* or graft)).tw,kf.
20. (1 OR 2 OR 3 OR 4 OR 5 OR 6 OR 7 OR 8 OR 9 OR 10 OR 11 OR 12 OR 13 OR 14 OR 15 OR 16 OR 17 OR 18 OR 19)
21. Exp Schizophrenia/
22. Exp Affective disorders, psychotic/
23. Exp Bipolar disorder/
24. (bipolar adj1 (disorder* or disease* or illness*)).tw,kf.
25. paranoid disorders/
26. Exp psychotic disorders/
27. schizo*.tw,kf.
28. (mani* adj3 depress*).tw,kf.
29. (psychotic* adj3 depress*).tw,kf.
30. (severe* adj3 affective*).tw,kf.
31. (severe* adj3 mental*).tw,kf.
32. (severe* adj3 depress*).tw,kf.
33. (psychos#s adj3 depress*).tw,kf.
34. (serious* adj3 affective*).tw,kf.
35. ‘serious mood*’.tw,kf.
36. (serious* adj3 mental*).tw,kf.
37. (serious* adj3 depress*).tw,kf.
38. ‘Severe mental*’.tw,kf.
39. Severe adj1 (mental* or depress*).tw,kf.
40. ‘Schizoaffective disorder’.tw,kf
41. Exp Antipsychotic Agents/
42. Exp Antimanic Agents/
43. Exp Psychotropic Drugs/
44. (21 OR 22 OR 23 OR 24 OR 25 OR 26 OR 27 OR 28 OR 29 OR 30 OR 31 OR 32 OR 33 OR 34 OR 35 OR 36 OR 37 OR 38 OR 39 OR 40 OR 41 OR 42 OR 43)
45. Exp epidemiologic studies/
46. Exp epidemiology/
47. epidemiolog*.tw,kf.
48. Exp prevalence/
49. prevalence.tw,kf.
50. Exp incidence/
51. incidence.tw,kf.
52. Exp Observational Study/
53. observational.tw,kf.
54. Longitudinal Studies/
55. longitudinal.tw,kf.
56. Case-Control Studies/
57. Exp Cross-Sectional Studies/
58. Exp Cohort Studies/
59. ‘cohort’.tw,kf.
60. Exp Risk/
61. Exp Risk Factors/
62. (45 OR 46 OR 47 OR 48 OR 49 OR 50 OR 51 OR 52 OR 53 OR 54 OR 55 OR 56 OR 57 OR 58 OR 59 OR 60 OR 61)
63. (20 AND 44 AND 62)

## References

1. Fiorillo A, Sartorius N. Mortality gap and physical comorbidity of people with severe mental disorders: the public health scandal. Annals of General Psychiatry. 2021;20(1):52. doi: 10.1186/s12991-021-00374-y.

2. Siddiqi N, Doran T, Prady SL, Taylor J. Closing the mortality gap for severe mental illness: Are we going in the right direction? British Journal of Psychiatry. 2017;211(3):130-1. Epub 2018/01/02. doi: 10.1192/bjp.bp.117.203026.

3. Hayes JF, Marston L, Walters K, King MB, Osborn DPJ. Mortality gap for people with bipolar disorder and schizophrenia: UK-based cohort study 2000-2014. Br J Psychiatry. 2017;211(3):175–81. Epub 20170706. doi: 10.1192/bjp.bp.117.202606. PubMed PMID: 28684403; PubMed Central PMCID: PMCPMC5579328.

4. Nielsen RE, Banner J, Jensen SE. Cardiovascular disease in patients with severe mental illness. Nature Reviews Cardiology. 2021;18(2):136–45. doi: 10.1038/s41569-020-00463-7.

5. Correll CU, Solmi M, Veronese N, Bortolato B, Rosson S, Santonastaso P, et al. Prevalence, incidence and mortality from cardiovascular disease in patients with pooled and specific severe mental illness: a large-scale meta-analysis of 3,211,768 patients and 113,383,368 controls. World Psychiatry. 2017;16(2):163–80. doi: 10.1002/wps.20420.

6. Vancampfort D, Correll CU, Galling B, Probst M, De Hert M, Ward PB, et al. Diabetes mellitus in people with schizophrenia, bipolar disorder and major depressive disorder: a systematic review and large scale meta-analysis. World Psychiatry. 2016;15(2):166–74. doi: 10.1002/wps.20309.

7. Carswell C, Cogley C, Bramham K, Chilcot J, Noble H, Siddiqi N. Chronic kidney disease and severe mental illness: a scoping review. Journal of Nephrology. 2023;36(6):1519–47. doi: 10.1007/s40620-023-01599-8.

8. Kalantar-Zadeh K, Jafar TH, Nitsch D, Neuen BL, Perkovic V. Chronic kidney disease. The Lancet. 2021;398(10302):786–802. doi: 10.1016/S0140-6736(21)00519-5.

9. Kovesdy CP. Epidemiology of chronic kidney disease: an update 2022. Kidney Int Suppl (2011). 2022;12(1):7–11. Epub 20220318. doi: 10.1016/j.kisu.2021.11.003. PubMed PMID: 35529086; PubMed Central PMCID: PMCPMC9073222.

10. Foreman KJ, Marquez N, Dolgert A, Fukutaki K, Fullman N, McGaughey M, et al. Forecasting life expectancy, years of life lost, and all-cause and cause-specific mortality for 250 causes of death: reference and alternative scenarios for 2016-40 for 195 countries and territories. Lancet. 2018;392(10159):2052–90. Epub 20181016. doi: 10.1016/s0140-6736(18)31694-5. PubMed PMID: 30340847; PubMed Central PMCID: PMCPMC6227505.

11. Francis A, Harhay MN, Ong ACM, Tummalapalli SL, Ortiz A, Fogo AB, et al. Chronic kidney disease and the global public health agenda: an international consensus. Nature Reviews Nephrology. 2024;20(7):473–85. doi: 10.1038/s41581-024-00820-6.

12. Grünfeld J-P, Rossier BC. Lithium nephrotoxicity revisited. Nature Reviews Nephrology. 2009;5(5):270–6. doi: 10.1038/nrneph.2009.43.

13. Gupta S, Khastgir U. Drug information update. Lithium and chronic kidney disease: debates and dilemmas. BJPsych Bull. 2017;41(4):216–20. doi: 10.1192/pb.bp.116.054031. PubMed PMID: 28811917; PubMed Central PMCID: PMCPMC5537577.

14. Højlund M, Lund LC, Herping JLE, Haastrup MB, Damkier P, Henriksen DP. Second-generation antipsychotics and the risk of chronic kidney disease: a population-based case-control study. BMJ Open. 2020;10(8):e038247. doi: 10.1136/bmjopen-2020-038247.

15. Abosi O, Lopes S, Schmitz S, Fiedorowicz JG. Cardiometabolic effects of psychotropic medications. Horm Mol Biol Clin Investig. 2018;36(1). Epub 20180110. doi: 10.1515/hmbci-2017-0065. PubMed PMID: 29320364; PubMed Central PMCID: PMCPMC6818518.

16. Ndumele CE, Rangaswami J, Chow SL, Neeland IJ, Tuttle KR, Khan SS, et al. Cardiovascular-Kidney-Metabolic Health: A Presidential Advisory From the American Heart Association. Circulation. 2023;148(20):1606–35. doi: 10.1161/CIR.0000000000001184.

17. Fu YC, Xu ZL, Zhao MY, Xu K. The Association Between Smoking and Renal Function in People Over 20 Years Old. Front Med (Lausanne). 2022;9:870278. Epub 20220603. doi: 10.3389/fmed.2022.870278. PubMed PMID: 35721101; PubMed Central PMCID: PMCPMC9205397.

18. Kuma A, Kato A. Lifestyle-Related Risk Factors for the Incidence and Progression of Chronic Kidney Disease in the Healthy Young and Middle-Aged Population. Nutrients. 2022;14(18). Epub 20220914. doi: 10.3390/nu14183787. PubMed PMID: 36145162; PubMed Central PMCID: PMCPMC9506421.

19. Dickerson F, Schroeder J, Katsafanas E, Khushalani S, Origoni AE, Savage C, et al. Cigarette Smoking by Patients With Serious Mental Illness, 1999–2016: An Increasing Disparity. Psychiatric Services. 2017;69(2):147–53. doi: 10.1176/appi.ps.201700118.

20. Vancampfort D, Firth J, Schuch FB, Rosenbaum S, Mugisha J, Hallgren M, et al. Sedentary behavior and physical activity levels in people with schizophrenia, bipolar disorder and major depressive disorder: a global systematic review and meta-analysis. World Psychiatry. 2017;16(3):308–15. doi: 10.1002/wps.20458.

21. Dipasquale S, Pariante CM, Dazzan P, Aguglia E, McGuire P, Mondelli V. The dietary pattern of patients with schizophrenia: A systematic review. Journal of Psychiatric Research. 2013;47(2):197–207. doi: 10.1016/j.jpsychires.2012.10.005.

22. Teasdale SB, Ward PB, Samaras K, Firth J, Stubbs B, Tripodi E, et al. Dietary intake of people with severe mental illness: systematic review and meta-analysis. The British Journal of Psychiatry. 2019;214(5):251–9. Epub 2019/02/20. doi: 10.1192/bjp.2019.20.

23. Okpechi IG, Caskey FJ, Gaipov A, Tannor EK, Noubiap JJ, Effa E, et al. Early Identification of CKD-A Scoping Review of the Global Populations. Kidney Int Rep. 2022;7(6):1341–53. Epub 20220406. doi: 10.1016/j.ekir.2022.03.031. PubMed PMID: 35685314; PubMed Central PMCID: PMCPMC9171699.

24. Neuen BL, Jun M, Wick J, Kotwal S, Badve SV, Jardine MJ, et al. Estimating the population-level impacts of improved uptake of SGLT2 inhibitors in patients with chronic kidney disease: a cross-sectional observational study using routinely collected Australian primary care data. The Lancet Regional Health – Western Pacific. 2024;43. doi: 10.1016/j.lanwpc.2023.100988.

25. Jankowski J, Floege J, Fliser D, Böhm M, Marx N. Cardiovascular Disease in Chronic Kidney Disease. Circulation. 2021;143(11):1157–72. doi: 10.1161/CIRCULATIONAHA.120.050686.

26. de Mooij LD, Kikkert M, Theunissen J, Beekman ATF, de Haan L, Duurkoop P, et al. Dying Too Soon: Excess Mortality in Severe Mental Illness. Front Psychiatry. 2019;10:855. Epub 20191206. doi: 10.3389/fpsyt.2019.00855. PubMed PMID: 31920734; PubMed Central PMCID: PMCPMC6918821.

27. Hsu Y-H, Cheng J-S, Ouyang W-C, Lin C-L, Huang C-T, Hsu C-C. Lower Incidence of End-Stage Renal Disease but Suboptimal Pre-Dialysis Renal Care in Schizophrenia: A 14-Year Nationwide Cohort Study. PLOS ONE. 2015;10(10):e0140510. doi: 10.1371/journal.pone.0140510.

28. Boyle SM, Fehr K, Deering C, Raza A, Harhay MN, Malat G, et al. Barriers to kidney transplant evaluation in HIV-positive patients with advanced kidney disease: A single-center study. Transplant Infectious Disease. 2020;22(2):e13253. doi: 10.1111/tid.13253.

29. Tzur Bitan D, Krieger I, Berkovitch A, Comaneshter D, Cohen A. Chronic kidney disease in adults with schizophrenia: A nationwide population-based study. General Hospital Psychiatry. 2019;58:1–6. doi: 10.1016/j.genhosppsych.2019.01.007.

30. Bayat S, Frimat L, Thilly N, Loos C, Briançon S, Kessler M. Medical and non-medical determinants of access to renal transplant waiting list in a French community-based network of care. Nephrology Dialysis Transplantation. 2006;21(10):2900–7. doi: 10.1093/ndt/gfl329.

31. Carswell CC, Joseph; Jacobs, Rowena; Siddiqi, Najma; Osborn, David. The prevalence of chronic kidney disease amongst people with severe mental illness: a systematic review and meta-analysis.: PROSPERO; 2024.

32. Moher D, Shamseer L, Clarke M, Ghersi D, Liberati A, Petticrew M, et al. Preferred reporting items for systematic review and meta-analysis protocols (PRISMA-P) 2015 statement. Systematic Reviews. 2015;4(1):1. doi: 10.1186/2046-4053-4-1.

33. Reilly S, Olier I, Planner C, Doran T, Reeves D, Ashcroft DM, et al. Inequalities in physical comorbidity: a longitudinal comparative cohort study of people with severe mental illness in the UK. BMJ Open. 2015;5(12):e009010. doi: 10.1136/bmjopen-2015-009010.

34. Osborn DPJ, Wright CA, Levy G, King MB, Deo R, Nazareth I. Relative risk of diabetes, dyslipidaemia, hypertension and the metabolic syndrome in people with severe mental illnesses: Systematic review and metaanalysis. BMC Psychiatry. 2008;8(1):84. doi: 10.1186/1471-244X-8-84.

35. Das-Munshi J, Chang C-K, Dutta R, Morgan C, Nazroo J, Stewart R, et al. Ethnicity and excess mortality in severe mental illness: a cohort study. The Lancet Psychiatry. 2017;4(5):389–99. doi: 10.1016/S2215-0366(17)30097-4.

36. Taylor J, Stubbs B, Hewitt C, Ajjan RA, Alderson SL, Gilbody S, et al. The Effectiveness of Pharmacological and Non-Pharmacological Interventions for Improving Glycaemic Control in Adults with Severe Mental Illness: A Systematic Review and Meta-Analysis. PLOS ONE. 2017;12(1):e0168549. doi: 10.1371/journal.pone.0168549.

37. Innovation. VH. Covidence systematic review software.

38. Munn Z, Moola S, Lisy K, Riitano D, Tufanaru C. Methodological guidance for systematic reviews of observational epidemiological studies reporting prevalence and cumulative incidence data. Int J Evid Based Healthc. 2015;13(3):147–53. doi: 10.1097/xeb.0000000000000054. PubMed PMID: 26317388.

39. Schünemann HB, Jan; Guyatt, Gordon; Oxman, Andrew. GRADE Handbook 2013. Available from: https://gdt.gradepro.org/app/handbook/handbook.html.

40. Edwards J, Hayden J, Asbridge M, Gregoire B, Magee K. Prevalence of low back pain in emergency settings: a systematic review and meta-analysis. BMC Musculoskeletal Disorders. 2017;18(1):143. doi: 10.1186/s12891-017-1511-7.

41. Borges Migliavaca C, Stein C, Colpani V, Barker TH, Munn Z, Falavigna M. How are systematic reviews of prevalence conducted? A methodological study. BMC Med Res Methodol. 2020;20(1):96. Epub 20200426. doi: 10.1186/s12874-020-00975-3. PubMed PMID: 32336279; PubMed Central PMCID: PMCPMC7184711.

42. Migliavaca CB, Stein C, Colpani V, Barker TH, Ziegelmann PK, Munn Z, et al. Meta-analysis of prevalence: statistic and how to deal with heterogeneity. Research Synthesis Methods. 2022;13(3):363–7. doi: 10.1002/jrsm.1547.

43. Munn Z, Moola S, Lisy K, Riitano D, Tufanaru C. Methodological guidance for systematic reviews of observational epidemiological studies reporting prevalence and cumulative incidence data. JBI Evidence Implementation. 2015;13(3).

44. Schoretsanitis G, de Filippis R, Brady BM, Homan P, Suppes T, Kane JM. Prevalence of impaired kidney function in patients with long-term lithium treatment: A systematic review and meta-analysis. Bipolar Disorders. 2022;24(3):264–74. doi: 10.1111/bdi.13154.

